# Improved RT-PCR SARS-Cov2 results interpretation by indirect determination of cut-off cycle threshold value

**DOI:** 10.1101/2020.11.20.20235390

**Authors:** Khelil Mohamed Mokhtar

## Abstract

Clinical laboratories of the developing world are overwhelmed with RT-PCR SARS-Cov2 testing demands. It is of paramount importance that each clinical laboratory use an appropriate cut-off value in the interpretation of SARS-Cov2 real-time RT–PCR results, which is specific to their laboratory performances as ISO 15189 recommendations stipulate. We applied an indirect statistical method to a large mixed data set of Ct values (ORF1ab and N) to estimate cut-off Ct value (∼32 cycles).we conclude that the use of indirect statistical approaches to estimate cut-off value in the interpretation of SARS-Cov2 real-time RT–PCR results may improve differential diagnosis of COVID-19 cases with low risk of infectivity, and may help to better estimates of the burden of COVID-19 disease.

---

Dear editor;

Clinical laboratories played a key role in controlling spread of SARS-COV2 virus by rapid diagnosis of COVID-19 cases which aid in rapid isolation of confirmed cases and close contacts, with social movement constrain (1). On 27 May 2020, WHO provided updated recommendations on the criteria for discharging patients from isolation. Patients were discharged 13 days after symptom onset for symptomatic cases and 10 days after positive test for SARS-Co2 for asymptomatic (2). These recommendations were based on evidences showing the low probability of virus culture in upper respiratory samples after 9-10 days after symptom onset (3-4). Only a few studies have studied how RT-PCR detection (i.e.: Cut-off cycle threshold “Ct”) relates to cultivable virus (4-5). However there was a general agreement that Ct values were strongly associated with the ability to recover infectious SARS-COV2 virus .Furthermore a cycle threshold cut-off 33-35 was associated with low infectivity. Clinical laboratories often use Ct cut-off provided by the RT-PCR kit manufacturers which are between 37 and 40 cycles. Ideally, each clinical laboratory should perform its own correlation between culture results and viral RNA load from patients’ samples, but the process is not widely available and require biosafety level 3 facilities. The aim of this study was to estimate a local cut-off value for SARS-Cov2 viral load by data-mining using intra-laboratory Ct values, which can be used as a surrogate for infectiousness. We started from the observation that distributions of ORF1ab and N Ct values in screening cases are typically bimodal. We hypothesized that the lower Ct value peak corresponds to patients with high infectivity, and the higher Ct value peak corresponds to patients for with low infectivity. We modelled the ORF1ab and N Ct values bimodal distributions with a finite-mixture model -alias maximum likelihood model- and estimated local Ct Cut-off values associated with SARS-Cov2 infectivity risk.

The dataset used in this study were generated from a retrospective, outpatients based COVID-19 results, in INSTITUT PASTEUR OF M’SILA, ALGERIA. Only screening results were included. SARS-CoV-2 RNA positivity was assessed by real-time reverse transcription-PCR targeting the ORF1ab and N genes, as previously described (7). We assumed that high and low infectivity populations follow a Gaussian distribution. The bimodal distribution of Ct values of ORF1an and N was modelled by the finite mixture model using an open source software package (mixtool) in the R programming language (8-9).For each distribution, parameters λ, μ1, σ1, μ2, σ2 were estimated by the maximum likelihood approach using the expectation maximization (EM) algorithm, the confidence intervals were calculated using the formula: 95% 95% CI = +/- 2.81* σ/√*n* as proposed by Boyd (10).

From the 13^th^ July to the 13th October 2020, a total of 3076 nasopharyngeal (NP) samples from outpatients (1230 female and 1846 male) were collected and examined for SARS-Cov-2 RNA presence. The mean (median) age of female was 54 years (55 years) and for male was 55 years (55 years). 2444 Ct values (N and ORF1ab gene) of BIOGERM™ RT-PCR kit were collected and analysed. From 2444 Ct values there were only 41 repeats, medians of Ct were 35.44 and 35.25 for ORF1ab and N gene respectively.

The finite mixture model was applied to fit two Gaussian distributions in the mixed data set (Ct values of ORF1ab and N =< 45) (Figure1.A and B).For ORF1ab datasets, the 2.5^th^ percentile of the right curve (in green) and the 97.5^th^ percentile of the left curve (in red) were 29.2 (95%CI: 29.01-29.5) and 32.3 (95%CI: 32-32.6) respectively. For N dataset, the 2.5th percentile of the right curve (in green) and the 97.5th percentile of the left curve (in red) were 27.9 (95% CI: (27.6-28.1) and 30.7 (95%CI: 30.5 - 30.9) respectively. Using the 2.5^th^ percentiles (∼29) and the 97.th percentiles (∼32) as cut-offs, the adjusted COVID-19 detection rates reduced significantly from 89% to 36% and 52% respectively.

**Figure1.**
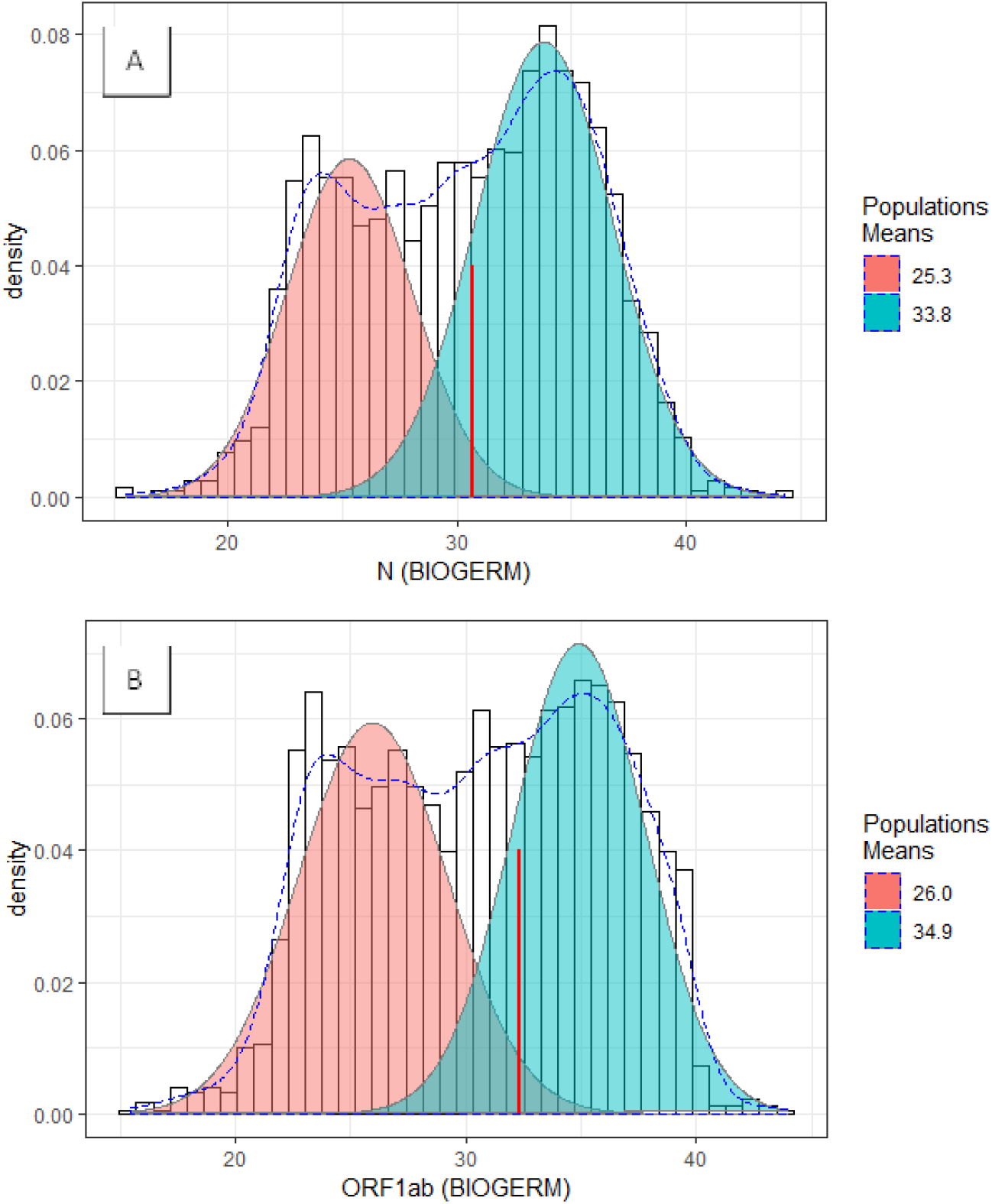
Maximum likelihood modeling of Ct values distributions for N gene (Figure 2A) and ORF1ab gene (BIOGERM™) (Figure 2B). The curves in the left (in red) represent high infectiousness population and the curves in the right (in green) represent low infectiousness population. Dashed blue curves display the mixed population. Red vertical segments represent the Cut-off Cycle thresholds.

To the best of our knowledge, cut-off cycle threshold value of SARS-cov2 RT-PCR has not been estimated previously by indirect statistical procedures .In this study, we propose an indirect approach to distinguish COVID-19 cases with high likelihood of infectiousness versus cases with low likelihood of infectiousness. Our method apply the finite-mixture model to local retrospectives dataset of ORF1ab and N Ct values. It was founded on the observation that the distribution of Ct values for ORF1ab and N is bimodal, and the assumption that the central part of the total distribution represents the low infectivity population (11). Our results show that the 97.5^th^ percentiles of the lower peaks (ORF1ab and N) which is (∼32 cycles) may represents the maximum diagnostic efficiency cut-off. Our findings have been demonstrated by two ways: (1) our model-based cut-off was different but close to that obtained by La Scola et al (33-34 cycles) and Singanayagam et al (35 cycles). This was expected, due to the differences in laboratory practices (i.e.; different sampling technique and medium, RNA extraction methods, method sensitivity).(2) After applying the estimated cut-off (Ct≤32) to our dataset, the proportion of COVID-19 positive cases decreased by 37% compared to 20% (Ct≤35).

We conclude that indirect statistical methods may allow the estimation of cut-off cycle threshold value of SARS-COV2 RT-PCR from a large dataset stored in laboratory information system. We suggest that each clinical laboratory conducting routine diagnosis of COVID-19, should determine its own cut-off depending on its own laboratory practices and analytical method performances. We argue that it may be necessary for some regional laboratories with similar patient’s characteristics using the same pre-analytical and analytical procedures to form a common data pool and calculate common cut-off Ct value, which could aid to improve our understanding of the burden of COVID-19 disease.

## Data Availability

No data availability

